# Exploring Factors Influencing Adherence to HIV Medication among patients using Community Pharmacies and Public Hospital: A Qualitative Study

**DOI:** 10.1101/2024.04.16.24305580

**Authors:** Odunayo Betty Olaleye, Olayinka Stephen Ilesanmi

**Affiliations:** Department of Public Health, Texila American University, Guyana, South America; Department of Community Medicine, College of Medicine, University of Ibadan, Oyo State, Nigeria

**Keywords:** ART adherence, HIV medication, Qualitative study

## Abstract

**Introduction:** HIV/AIDS is a major global health emergency as it is estimated that close to 40 million people live with HIV/AIDS, among which the majority are adults. Despite the availability of ARVs at community, secondary, and tertiary health facilities in Nigeria, adherence rates remain low. This study conducted in Ibadan, Nigeria, investigated factors influencing adherence to antiretroviral therapy (ART) among HIV patients.

**Methods:** The research employed a cross-sectional approach, utilizing qualitative techniques, including in-depth interviews (IDI) with patients and key informant interviews (KII) with healthcare providers and pharmacists. The target population consisted of HIV patients receiving ART at the Community Pharmacies and Public Hospital University in Ibadan. Data saturation determined sample sizes for the interviews.

**Results:** In the in-depth interviews, two primary themes emerged from the findings: “Socio-economic factors influencing adherence” and “Factors related to the process of care.” Socio-economic factors encompassed issues such as transportation costs, societal pressure, food costs, advice from third parties, and religious reasons. These factors were further categorized into economic reasons and societal pressure/misinformation, shedding light on patients’ challenges and choices. Factors related to the process of care included the cost of medication, side effects, drug-drug interactions, the quantity and duration of drug use, and discontinuation after feeling better.

Key informant interviews with healthcare providers and pharmacists revealed insights into compliance by gender, financial constraints, and treatment-related factors such as side effects, drug interactions, and medication volume. Socio Cultural and religious factors, including advice from third parties, stigmatization, disclosure of status to partners, social support, and religious beliefs, played significant roles in influencing patients’ adherence.

**Conclusion:** These findings shed light on the complex interplay of factors affecting ART adherence and highlight the importance of considering economic, social, and medical aspects when designing interventions to improve adherence in HIV care. The study’s findings hold implications for healthcare programs and interventions aimed at enhancing the quality of care and support for HIV patients, particularly in resource-limited settings like Southwestern Nigeria.

## Introduction

HIV/AIDS stands as a critical global health emergency, impacting diverse populations and precipitating a substantial toll of suffering and mortality across the world (1). It affects a wide spectrum of individuals, spanning geographical regions, countries, age groups, and demographics, encompassing adults, children, and pregnant women. Globally, almost 40 million people live with HIV/AIDS, with a predominant majority being adults, while a minority comprises children (2). A 2019 report by UNAIDS and the National Agency for the Control of AIDS (NACA) estimated that Nigeria is home to 1.9 million people living with HIV (3).

Adherence to ART treatment (complying strictly to medication regimen instructions) is a critical component of effective management of HIV/AIDS (4). Poor adherence is a serious public health concern and was reported as one of the ways of introducing complication to HIV/AIDS as it can lead to the viral resistance thereby leading to failure of cheap first-line treatment regimens and increase the spread of multidrug resistant forms of the virus (3). Effective treatment of HIV hinges on total adherence as the infection is a peculiar one unlike many other diseases, therefore it is vital that PLHIV strictly adhere to their treatment plans by taking all the doses of their drug regimen to prevent resistance and to improve their chances of survival and this is possible if they understand the level of adherences that is expected of them and by trying to overcome the factors that can lead to default from care.

Recent studies conducted in Nigeria have reported an adherence rate of 92.6% among HIV patients, with multiple factors influencing both adherence to treatment and disengagement from care. These factors span socioeconomic, demographic, and healthcare-related dimensions, revealing a complex interplay of influences (4,5). Despite numerous studies reporting high adherence rates to HIV care, they often neglect to address the rate of disengagement from care, which is associated with significant morbidity and mortality among both ART and pre-ART patients due to non-adherence (6). A follow-up study on HIV-ART care revealed that 37% of individuals living with HIV in Nigeria, who had initiated care, eventually disengaged from it. Moreover, disengagement from care correlated with factors such as decreasing age, pre-ART status, HIV stage 1 & IV, and baseline CD4 counts above 400 cells/mm^3^ (7).

The aim of the study is to investigate and understand the factors that influence patients’ adherence to HIV treatment plans. The study aims to explore and identify these factors through in-depth interviews (IDI) and key informant interviews (KII).

## Methods

### Study Area and Population

The study was conducted in Ibadan, the capital of Oyo State, Southwestern Nigeria, located at latitude 7.3775° N and longitude 3.9470° E. Ibadan is recognized as the largest indigenous city in Africa and was historically the administrative center of the Western Region during British colonial rule. (8)

The target population for this study included HIV patients receiving antiretroviral therapy (ART) at the University College Hospital (UCH), Ibadan, who had chosen to receive their medication refills at either Eleta Hospital, a secondary facility, or their preferred community pharmacies. The UCH ART clinic had approximately 6,339 registered patients, while there were 24 community pharmacies acting as refill or pickup centers, each catering to approximately 60 patients.

### Study Design

This study adopted a cross-sectional approach with qualitative techniques. It encompassed facility-based and community-based cross-sectional surveys and utilized In-depth interview guides (IDI) and Key informant interview (KII) guides. The study design was selected to explore patient and healthcare provider experiences, with a focus on documenting factors influencing ART adherence. In-depth interview (IDI) was used to document the perceived effectiveness of the two refill models from the patients attending each of them and the respective barriers and facilitating factors while Key informant interview (KII) guide was used to collect data from the community pharmacies and health workers on the perceived effectiveness of the two refill models. The interview was conducted in either Yoruba or English as preferred by the respondents and trained research assistants were equipped to carry out the task accordingly. The final data was translated to English language before data analysis to ensure uniformity in the analysis.

### Sample Size determination for Qualitative Study

Data saturation criterion was used to determine the sample size. A priori of thirteen (13) interviews was set for each of the facilities for the in-depth interview guide since a previous qualitative study conducted on a qualitative study of persons who are 100% adherence to antiretroviral therapy (Lewis et al., 2006). In accordance with the qualitative descriptive approach, the criterion of saturation (that is, situations whereby no new information surfaces with new cases). Data saturation criterion was used to determine the sample size

For the key informant interview (KII) guide, 30 health workers and 36 pharmacists were interviewed in the secondary based facilities, while 25 health workers and 36 pharmacists were interviewed in the community-based facilities.

### Inclusion and Exclusion Criteria

Patients aged 18-80 years of age at ART initiation and that commenced ART at least six months prior to interview and community pharmacies who had been providing ARV refill services for more than six months were included in the study. Also, healthcare providers like public health nurses, pharmacists and other personnel in the facility that attend to HIV-patients or give ARV refills and who consented to participate in the study were included in the study.

Patients aged 18-80 years of age at ART initiation and commenced ART at least six months prior to interview but have other HIV unrelated complications and staff working at the clinic for less than 3 months prior to the study were excluded from the study.

### Data Collection Procedure

All interviews strictly follow the IDI and KII guide within the duration of about one hour to one hour thirty minutes during which the participants were asked prompting questions from the guide. Other issues raised by respondents during the interviews were used as cues for additional prompting questions and for probing for deeper meanings to the whole study’s research questions. For IDI, the interviews were conducted with eligible ART recipients at the clinic by two research assistants simultaneously depending on the frequency of the participants. For KII, the interviews were conducted within the pharmacies’ premises and hospital premises and in participants’ area of choosing during the office working hours between 8 am-4 pm to show consideration for their busy schedules.

The recruitment period for this study is from 8^th^ July,2021 to 5^th^ October 2021.

### Qualitative Data Management

All the recorded data from the interviews conducted among the respondents were transcribed word for word after which the researcher typeset and read it severally to get familiar with the data. This is necessary as it will help the researcher to obtain a sense of totality during which significant statements will be underlined and extracted. Meanings of significant statements and sentences that have similar characteristics were labelled by the researcher. Different parts of the text that contained significant statements were marked with appropriate labels for the use in the narratives and quotations that were presented in the result section. The data was analysed with the use of atlas.ti software version 8.

### Validity and Reliability of instruments

There was an extensive review of literature to ensure appropriate content and face validity for the record review guide that was used to harvest the adherence rate.

### Ethical Statement

The study obtained ethical approval from Oyo State Ministry of Health Research Ethics Committee. A consent form was given to the participants to obtain their formal consent, which they then signed. The participants were informed that their participation was voluntary and that they would not face any consequences for choosing not to participate.

The data collected were strictly confidential and used only for research purposes. All the research assistants underwent training to learn how to maintain confidentiality of information. There was little or no risk to the respondents because the study did not utilize any invasive technique. Therefore, no physical harm was inflicted on study participants because they participated in this study.

## Results

### A In-depth interview (IDI)

The findings from the qualitative study identified two themes which were the factors influencing patients’ adherence to treatment plans and they are “Socio-economic factors influencing adherence” and “Factors related with process of care”.

#### Theme 1: Socio-economic factors influencing adherence to HIV medication

The patients from the two facilities had similar responses to the questions that were asked as some expressed that inability to cope with transportation costs, undue pressure from what people will say at their workplace when they see them using the drugs, food cost, advice from third parties and religious reasons were those factors influencing their adherence to treatment plans. These factors were further split into two sub themes to identify those related to “Economic reasons’’ and those factors related to “Societal pressure and misinformation”.

#### Sub-theme 1: Economic factors

Some of the patients were very specific about the factors that affected their inability to keep up with appointments or with the drug usage plan that was prescribed from the pick-up centres. These factors had to do financial and economic challenges as it was presented in the quotes presented below:

*“The money that I am spending from my house to this place is much and sometimes, when I don’t have it and the day that I was asked to come, I will wait till the day I will get money*.*”* ((A female, businesswoman), Secondary facility pick-up centre)

*“My place of work is far from here, though, I like it like that, and I can come, and no one will know where I go or come from and I cannot use it at work too, so I use it at night. My house is far too, and I used to have challenges with transport fare…It was easy to get drugs to use during COVID-19 because we were given 3-months drugs and I had extra before, then I coped with it during COVID -19*.*”* ((A woman between age 26-30 years old), Secondary facility pick-up centre)

#### Sub-theme 2: Societal pressure and misinformation

Most of the patients that reported other reasons apart from economic reasons, explained that they missed their appointments or stopped taking their medications because of the advice given by their friends and families and religious fathers. Although they returned to care after the viral load increased and were hospitalized, however, the effects of the advice and assurance of a healing miracle made them stop at that point. The quotes below were extracted from direct responses from the interviews:

*“Yes, that was before, I don’t use it when I go to vigils because my pastor must not see it, he has prayed for me and told me that my sickness will not come back. And when I don’t use the drugs, I usually don’t feel the effect until the last time when I was seriously sick. Now, no one teaches me to use the drugs, pastor, or no pastor, I am the only one that knows what is wrong with me”* ((A female Student between age 21-25 years old) Pharmacy facility pick-up centre).

*“The only time I ever listened to my mother that it was my half-sisters that were responsible for my predicament, she took me to pray at one mountain church and the prophet told me to stop using the drugs if I have faith that I will get well. I stopped and I was sick…(smile) you know the rest of the story*…*here I am*.*”* ((A woman between age 26-30 years old) Pharmacy facility pick-up centre).

*“Till now, my husband is always complaining that the drugs are too much for the 5 years that I have started, but I never listened to him, he and my children don’t have it and I am taking the drugs to make it remain like that*.*”* ((A businesswoman) Pharmacy Pick-up Centre).

#### Theme 2: “Factors related with process of care”

Some of the participants interviewed in this study also mentioned several factors that are associated with the reasons why they default from the care and these factors are related to what happens to them when they use drugs and how they feel about the drugs (frequency and volume). All the factors were titled as factors related to the process of care and included cost of medication, cost of medication, side effects of medication, advice from a third party, number of drugs to use. The responses from the lived experience that were recounted by the patients were presented below as quotes to support the appropriateness of the themes identified within the data:

*“Any time that I don’t have food to eat, I dare not use the drug, because I will feel very tired and won’t be able to go and work to see money to take care of myself*.*”* ((A businessman), Secondary facility pick-up centre).

*“Hmmmm, that was a very long time ago (When asked if he stopped taking the medication), because this is my eight years that I have been coming here, the money was not my problem, even then, I used to give some people money when I met them, and they are complaining about the money we used to pay then for blood test. At the early stage, when I noticed that I was reducing in weight, (If you look at me, you will see that I am fat small (The interviewee looks heavy in weight and tall)), and I was thinking, maybe it’s the effects of the drugs, I stopped, but I noticed that it was even worse and I nearly lost my eyesight, I came to complain, they test my blood and advised me to continue. That was the only time, and it was a long time ago. Now, my results are always good*.*”* ((A businessman between age 46-50 years old), Secondary facility pick-up Centre).

### B Key Informant Interview (KII)

Compliance by Gender:

*Patient (Pharm 1): “Women are like 70% and men 30%. Women take things more seriously, and their compliance level is high*.*”*

*patient (Pharm 2): “Usually, we have more women than men*.*”*

*patient (Pharm 3): “Male is like 30% and female 70%*.*”*

*patient (Pharm 4): “I don’t have the percentage, but the females are more than men*.*” seriously, and their compliance level is high.”*

*patient (Pharm 6): “We don’t have an equal number of men and women. Our clients are mostly women, 80%*.*”*

*patient (Pharm 7): “60% female and 40% male*.*”*

Factors Responsible for Default from Care

Theme One: Finance

Sub-theme One: Cost of Medication

*Health Worker (Pharm 1): “The cost of medication can bring about default. When you recommend drugs to patients and they can’t afford it, they will default*.*”*

*Health Worker (Pharm 9): “Finance can cause default. When healthcare is not accessible to them*…*”*

Sub-theme Two: Cost of Transportation

*Health Worker (Pharm 3): “They sometimes say some of them are sick or complain about transport fare*.*”*

*Health Worker (Pharm 5): “Then for some, not being able to transport themselves down to the pick-up centre*.*”*

*Health Worker (Pharm 6): “A lot of them have financial challenges and some of them don’t stay in Ibadan. We have patients that travel from the north to come and pick up their drugs in Ibadan” Pharm 10”If they relocate or are not around. Pick-up becomes difficult*.*”*

*Health Worker (Pharm 7): “Change of environment”*

Theme two: Treatment-related factors

Sub-theme One: Side effect of Medication

*Health Worker (Pharm 1): “Side effects of medication can cause default. When the patient cannot manage the side effects, they default*.*”*

*Health Worker (Pharm 5): “*…*some bad side effects that will make them stop*.*”*

*Health Worker (Pharm 5): “*…*the clients that have been decentralized to the community pharmacy are already stable in medication*.*”*

*Health Worker (Pharm 5): “Depending on the anti-retroviral, some people have insomnia, some have bad dreams depending on the ARV they are placed on*.*”*

*Health Worker (Pharm 9): “Reactions to drugs can cause a default like sleeping disorder*.*”*

*Health Worker (Pharm 1): “When there is drug-drug interaction, the manifestation will come out as a side effect to the patient*.*”*

Sub-theme two: Drug-Drug Interaction

*Health Worker (Pharm 1): “When there is drug-drug interaction, the manifestation will come out as a side effect to the patient”*.

*Health Worker (Pharm 2): “Drug-drug interaction can happen with herbs and there is drug-food interaction*.*”*

*Health Worker (Pharm 3): “Drug-drug interaction is a particular drug interacting with another, affecting the absorption and effect of the drug*.*”*

*Health Worker (Pharm 5): ““Drug to drug interaction can give some side effects that they don’t like so they might just discontinue and that’s why at times you must advise them on time. some even use herbal medication, so they can have some bad side effect that will make them stop,”*

*Health Worker (Pharm 7): “Drug to drug interaction can be a major problem*… *Maybe the patients are on antibiotics*.*”*

*Health Worker (Pharm 9): “When there is drug-drug interaction, the manifestation will come out as a side effect to the patient*.*”*

*Health Worker (Pharm 10): “For the drug-to-drug interaction, the healthcare provider is supposed to take the patient’s medical history*… *and measure the reactions of the drugs*.*”*

Sub-theme Three: Quantity of drug to use.

*Health Worker (Pharm 2): “If there are so many drugs to use, it happens that there is a stronger molecule of drug given to the patient*.*”*

Sub-theme Four: Duration of treatment

*Health Worker (Pharm 3): “Most of the time the patients get tired of taking the drugs because they have been on drugs for more than 6 years”*.

Subtheme Five: Discontinuation of medications after feeling better.

*Health Worker (Pharm 4): “They feel they are feeling well and some of them may discontinue, after suppression of the viral load”*.

Subtheme Six: Other factors (Physiological conditions, e.g., pregnancy, and availability of medications)

*Health Worker (Pharm 8): “Other factors include breastfeeding, and drug availability”*.

Theme Three: Socio Cultural and religious factors

Sub-theme One: Advice from third party

*Health Worker (Pharm 8): “Also, wrong advice from a third party*.*”*

*Health Worker (Pharm 10): “For the defaulters all we can do is to encourage them and talk out about their challenges and all lie to them after all encouragement even if they may still not take the drugs maybe their friends have introduced them to some herbal medication”*.

Subtheme Two: Stigmatization

*Health Worker (Pharm 8): “Stigmatization is actually the major issue because to open up can be difficult for them, some of them lie that they came for someone’s pick up”*.

*Health Worker (Pharm 4): “Another thing is stigmatization when they don’t want people to question them maybe because of where they are “*

*Health Worker (Pharm 5): “Stigmatization, sometimes when they come for their pickup, if they see wing thinking they know what they are coming for”*.

*Health Worker (Pharm 6): “Another is if they are not sure their privacy is guaranteed*.*”*

Sub-theme Three: Disclosure of Status to Partner

*Health Worker (Pharm 5): “there are lots of patient here that have not disclosed their status their partner, sometimes when I ask them that did u take it when you were supposed to take it, they will say that they can’t take it at that time that they were busy but I don’t ask further questions knowing fully well that they may not want to disclose it to their partner about the medication”*

Sub-theme Four: Social support

*Health Worker (Pharm 10): “Love and support from friends and family”*.

*Health Worker (Pharm 10): “Support from their loved ones can also be a challenge. For patient that has HIV the love they get from their loves one goes a long way”*

Subtheme Five: Religious beliefs

*Health Worker (Pharm 3): “Religion too can be a factor”*.

## Discussion

Socio-economic factors influencing adherence to HIV medication.

The qualitative study revealed that socio-economic factors play a significant role in influencing adherence to HIV medication among patients in both community pharmacies and the secondary facility. These factors include economic challenges and societal pressure/misinformation.

Economic factors emerged as a barrier to adherence, with patients struggling to cope with transportation costs and the financial burden of accessing HIV medications. This finding aligns with previous research highlighting the impact of economic constraints on medication adherence among people living with HIV (PLWH) (9). High medication costs, transportation expenses, and the financial burden of managing a chronic illness can hinder patients’ ability to consistently adhere to their treatment plans.

Societal pressure and misinformation were also identified as influential factors. Patients reported facing undue pressure from friends, family members, and religious leaders to discontinue their HIV medications. This finding is consistent with previous studies that have documented the influence of cultural and religious beliefs on treatment adherence among PLWH (10). Misguided advice from religious figures, promising healing without medication, can undermine adherence efforts and lead to negative health outcomes.

The implications of these findings are significant for healthcare providers, policymakers, and researchers working to improve adherence to HIV medication. Understanding the socio-economic factors that impact adherence allows for the development of targeted interventions to address these challenges. To address economic barriers, strategies such as financial assistance programs, insurance coverage for medications, and transportation support can be implemented to alleviate the financial burden on patients (11). These interventions can enhance medication affordability and accessibility, promoting adherence to HIV treatment.

Furthermore, interventions aimed at addressing societal pressure and misinformation should focus on education and awareness campaigns. Collaborating with religious leaders and community influencers to disseminate accurate information about HIV treatment can help dispel myths and misconceptions surrounding medication use (12). Support groups and counselling services can also provide a safe space for patients to discuss their concerns and receive social support, countering the negative influences they may encounter.

### Factors Related to the Process of Care

This theme highlights the factors that influence patients’ decisions to default from HIV care and treatment. Participants mentioned several aspects related to the process of care, including the cost of medication, side effects of medication, advice from third parties, and the number of drugs to use.

The cost of HIV medication emerged as a significant factor influencing adherence to treatment. Some patients reported that they would skip medication doses when they couldn’t afford food to take the medication with. This indicates a direct link between the economic situation of the patients and their ability to adhere to the prescribed treatment plan. Previous studies have also highlighted the impact of medication costs on adherence among PLWH (13). Reducing the financial burden of medication can improve adherence rates and overall treatment outcomes.

Patients’ experiences with medication side effects were another critical factor influencing their adherence. Concerns about potential adverse effects led some individuals to stop taking their medications, while others experienced improvements over time and continued their treatment. This finding aligns with previous research highlighting the importance of managing medication side effects to enhance adherence (14). Healthcare providers should proactively address side effects, provide appropriate counselling, and offer alternative treatment options if necessary.

Patients’ decisions to discontinue medication were also influenced by advice from friends, family members, and religious leaders. Some individuals stopped taking their medications based on misguided beliefs or the assurance of healing without the need for medication. This highlights the significance of social influences on treatment adherence. Similar findings have been reported in other studies, emphasizing the need for patient education, and involving support networks in adherence-promoting interventions (15).

The complexity of the treatment regimen, particularly the number of drugs to be taken, was mentioned as a factor contributing to non-adherence. Patients with more complicated regimens may find it challenging to adhere to multiple medications at different times of the day. Simplifying treatment regimens and providing clear instructions can aid in improving adherence (16).

The findings of this theme hold several implications for healthcare providers and policymakers aiming to improve adherence to HIV medication. Understanding the factors related to the process of care can guide the development of targeted interventions to address these barriers effectively. Healthcare providers should prioritize patient-centred care, considering individual patient needs and circumstances. Addressing financial barriers through the provision of affordable or subsidized medication can help reduce the burden of treatment costs on patients.

Furthermore, healthcare professionals should proactively discuss potential side effects of medications with patients and provide support and guidance in managing these effects. Patient education programs can play a crucial role in dispelling misinformation and misconceptions about HIV treatment, particularly those stemming from advice received from third parties.

Simplification of treatment regimens and the use of combination therapies can also enhance adherence. Healthcare providers should work collaboratively with patients to develop personalized treatment plans that are manageable and align with the patients’ daily routines and lifestyles. Understanding the factors related to the process of care that influence adherence to HIV medication is essential in developing effective strategies to improve treatment adherence. By addressing cost-related challenges, managing side effects, countering misinformation, and simplifying treatment regimens, healthcare systems can enhance adherence rates and contribute to better health outcomes for people living with HIV.

### Factors Responsible for Default from Care

This study investigated the factors associated with patients’ adherence to HIV care and treatment, as well as the reasons for treatment default. The data were collected through interviews with pharmacists who interacted closely with the patients, allowing them to provide valuable insights into the patients’ behaviours’ and challenges related to medication adherence.

The cost of medication and transportation emerged as significant barriers to adherence. When patients couldn’t afford their medications or faced financial challenges related to transportation, they were more likely to default from care. These findings are consistent with previous research that highlights the impact of financial constraints on medication adherence among people living with HIV (17).

A prominent sub-theme under treatment-related factors was the side effects of medication. Patients who experienced severe side effects, such as sleeping disorders or bad dreams, were more likely to discontinue their treatment. Drug-drug interactions were also mentioned as potential causes of side effects, leading to non-adherence. Similar findings have been reported in other studies, emphasizing the need for careful monitoring and management of medication side effects to improve adherence (18,19).

Another sub-theme in this category was the quantity and duration of treatment. Some patients may feel overwhelmed by the number of drugs they need to take, especially if there are stronger or multiple medications involved. Additionally, long-term treatment can lead to treatment fatigue, where patients may become tired of adhering to their medications over an extended period. Addressing these factors requires patient education and counselling on the importance of continuous treatment (20).

Stigmatization was a significant socio-cultural factor affecting adherence. Some patients felt uncomfortable taking their medications in the presence of others, especially when there was a fear of being judged or discriminated against due to their HIV status. Stigma has been widely recognized as a barrier to HIV care and treatment, and efforts to reduce stigma can positively impact adherence rates (21).

Another sub-theme was the influence of advice from third parties, which could sometimes be misguided or encourage patients to seek alternative treatments, such as herbal medications. This highlights the importance of involving support networks and providing accurate information about HIV treatment and its benefits (22).

Social support and religious beliefs were also mentioned as factors influencing adherence. Patients who received love and support from friends and family were more likely to adhere to their treatment. Additionally, religious beliefs can influence treatment decisions, and some patients may rely on faith-based practices rather than solely on medical interventions.

Healthcare providers should consider cultural and religious beliefs when providing counselling and support (23,24).

## Abbreviations

AIDS: Acquired Immune Deficiency Disease Syndrome
ART: Antiretroviral treatment
HIV: Human Immunodeficiency Virus

## Statements

### Statement of Ethics

#### Study approval statement

This study protocol was reviewed and approved by the Ministry of Health, department of planning, research, and statistics division. (UI/UCH Ethics Review Committee),)(approval number (AD 13/479/4045))

#### Consent to participate statement

Written informed consent was obtained from participants to participate in the study.

### Author Contributions

Both authors contributed to the writing of this manuscript for publication.

Odunayo Betty Olaleye analysed the data and prepared the manuscript.

Olayinka Stephen Ilesanmi review and edit.

Both authors approved the final manuscript.

### Data Availability Statement

All data generated or analysed during this study are included in this article. further enquiries can be directed to the corresponding author.

